# A methodological blueprint to identify COVID-19 vulnerable locales by socioeconomic factors, developed using South Korean data

**DOI:** 10.1101/2020.10.26.20218842

**Authors:** Bayarmagnai Weinstein, Alan R. da Silva, Dimitrios E. Kouzoukas, Tanima Bose, Gwang-Jin Kim, Paola A. Correa, Santhi Pondugula, Jihoo Kim, David O. Carpenter

**Affiliations:** University at Albany, Rensselaer, New York, USA; University of Brasília, Brasília, Brazil; Edward Hines Jr. VA Hospital, Hines, Illinois, USA; Loyola University Chicago, Maywood, Illinois, USA; Ludwig-Maximilian University of Munich, Planegg-Martinsried, Germany; University of Freiburg, Freiburg, Germany; Howard Hughes Medical Institute, Ashburn, Virginia, USA; University of Florida, Florida, USA; Hanyang University, Seoul, South Korea; Harvard University, Boston, Massachusetts, USA

**Keywords:** COVID-19, Pandemics, Socioeconomic Factors, Spatial Regression, South Korea

## Abstract

COVID-19 has more severely impacted socioeconomically (SES) disadvantaged populations. Lack of SES measurements and inaccurately identifying high-risk locales can hamper COVID-19 mitigation efforts. Using South Korean COVID-19 incidence data (January 20 through July 1, 2020) and established social theoretical approaches, we identified COVID-19-specific SES factors. Principal component analysis created composite indexes for each SES factor, while Geographically Weighted Negative Binomial Regressions mapped a continuous surface of COVID-19 risk for South Korea. High area morbidity, risky health behaviours, crowding, and population mobility elevated area risk for COVID-19, while improved social distancing, healthcare access, and education decreased it. Our results indicated that falling SES-related COVID-19 risks and spatial shift patterns over three consecutive time periods reflected the implementation of reportedly effective public health interventions. While validating earlier studies, this study introduced a methodological blueprint for precision targeting of high-risk locales that is globally applicable for COVID-19 and future pandemics.

## Introduction

COVID-19 corroborated insights gained from SARS, H1N1 influenza, and MERS pandemics that more severely affect socioeconomically (SES)-disadvantaged populations^1^. Area-health and SES lead to differential incidence and mortality as they define the disease agent exposure extent, susceptibility to contracting disease once exposed, and disease severity^2^. Although existing studies provided COVID-19 risk factors, none identified COVID-19-vulnerable locales associated with SES factors with acceptable generalizability and methodological capacity.

Current studies use isolated arbitrary SES variables based on researchers’ preferences. These lack conceptual clarity, and thereby fail to truly uncover how SES affects health outcomes. Consequently, a lack of harmony (reduced external validity) exists among the studies conducted across national and sub-national settings. This lack impedes rapid response and effective control measures, especially for developing countries relying on resource-rich countries for guidance. To address this, we integrated Coleman’s Social Theory and Blumenshine’s mechanistic framework (Figure 1), which permits a universal SES definition and SES indicator selection mechanistically/causally relevant to the health outcome.

**Figure 1.**
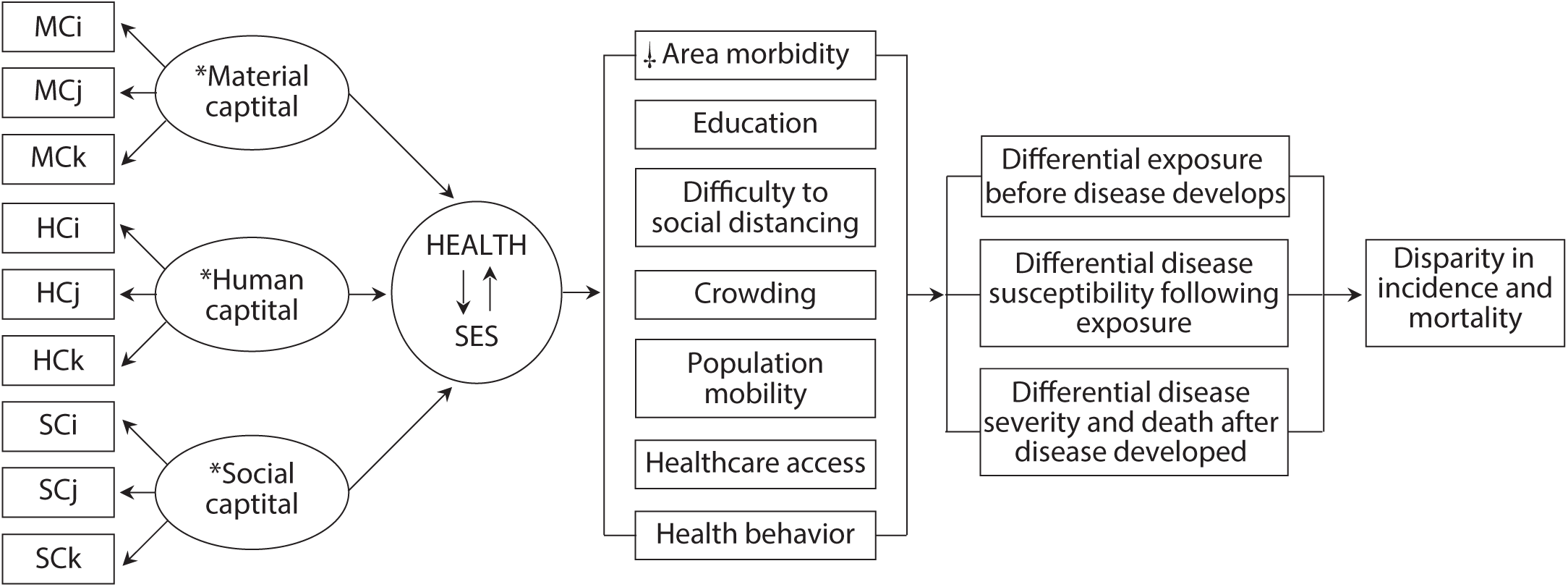
Conceptual model of the causal relationship between the SARS-CoV-2 and area health/SE determinants. Subscripts (i, j, k): Number of variables used from the data sources. Data sources: Korean Community Health Survey by the KCDC, Health Insurance Statistics by the National Health Insurance Services, Disability Status by the Ministry of Health and Welfare, Death Cause Statistics by the National Statistics Agency, Korean Census Bureau, Internal Migration Statistics by the Statistics Korea, and the State of Urban Planning Report by the Ministry of Land, Infrastructure, Transport, and Tourism. *Material, Human and Social Capital refers to latent structural components of the area health and SES determinants Health and SES connected by arrows indicate the integration of area health and SES as a composite exposure in this study, hereinafter, denoted as a health/SE. □Area-health/SE themes identified relevant to COVID-19 based on the person and population-level literature. Modified images from source^2, 33^.

SES data span an expanse of highly intercorrelated variables such as *healthcare utility rate, insurance coverage rate*, and *unmet healthcare needs*, each used interchangeably as a healthcare access measure. A composite measure approach can summarize the multi-dimensional information of individual SES variables while addressing the inadequacy of traditional statistics in simultaneously handling multiple highly intercorrelated variables. A commonly-used index, area-deprivation index (ADI)^3^, can adjust models to SES as a confounder, but not when SES is the exposure. Since disease risk is determined by multiple SES factors, understanding SES-related disease risk requires SES concept-specific data. Therefore, we recommend causally-important multiple composite indexes, as these allow researchers to delineate the mechanism through which SES theme(s) increase the COVID-19 risk^4^.

Thus far, only one study explored geographic methodology with a multi-scale Geographically Weighted Regression (GWR) accounting for selected SES variables (median household income, income inequality, percentage of nurse practitioners, and black female population) in the US^5^. Since multi-scale GWR doesn’t fit a beta distribution typical for infectious disease rates^6^, we recommend Geographically Weighted Negative Binomial Regression (GWNBR) to improve accuracy. This directly takes discrete count data without further transformation, and is robust to overdispersion, spatial/temporal clustering and false-positives^7, 8^.

Globally, the COVID-19 pandemic emerged in waves with country-specific mitigation strategies producing sharp declines. To improve public health interventions by precision targeting of high-risk locales, this study identified key SES risk determinants and their geographic distribution. We chose South Korean COVID-19 incidence data because it presented extremely high overdispersion and spatial clustering, being more complex than typical infectious disease data. South Korea, as an extreme case scenario, allowed us to check our framework’s functionality. Our study’s goals were to 1) provide methodological guidance for identifying COVID-19-vulnerable locales associated with SES factors, and 2) operationalize a framework using South Korean data to demonstrate its value and in the interpretation of the results.

## Methods

### Study design and population

We used COVID-19 incidence data from January 20 through July 1, 2020, released from the Korea Centers for Disease Control and Prevention (KCDC)^9^ and prepared by the DS4C project^10^. Analytical data consisted of 11 811 COVID-19 cases aggregated by 250 districts (Table S1) aligned to SAS’s South Korean geographic matrix. Since data was unavailable for Daegu’s subparts, we estimated the incidence from KCDC’s press release cluster reports.

### Conceptual model

Figure 1 shows the Coleman-Blumenshine Framework (CBF) refined approach, based on Coleman’s Social Theory and Blumenshine’s mechanistic framework^2, 11^. The model defines SES as a function of social and human capitals^11, 12^ and emphasizes pathways by how SES indicators differentially increase SARS-CoV-2 exposure and susceptibility to developing COVID-19^2^. Based on the CBF model and COVID-19 risk factors literature^13-17^, we identified seven area-level health and SES factors that determined the SARS-CoV-2 exposure level and the likelihood of developing COVID-19 after exposure.

### SES measurement

All SES related data sources were retrieved from the Korean Statistical Information Service’s (KOSIS) online data archive^18^. Table S3 presents the reviewed data sources used for SES measurement. Table 1 shows 24 data items out of 124 candidates relevant to the seven health/SE areas. We used an independent variable proxy for *education*, and by Principal Component Analysis (PCA) created six thematic composite indices: *healthcare access, health behaviour, crowding, area morbidity, education, difficulty to social distancing*, and *population mobility*. Factors were computed as linear combinations of the original variables selected for each health/SE theme. We used the first component scores^19^ in calculating the composite scores since they explained the largest data variation. Then we computed each variable’s weight by dividing each factor score by the sum of all variable factor scores as:

**Table 1.**
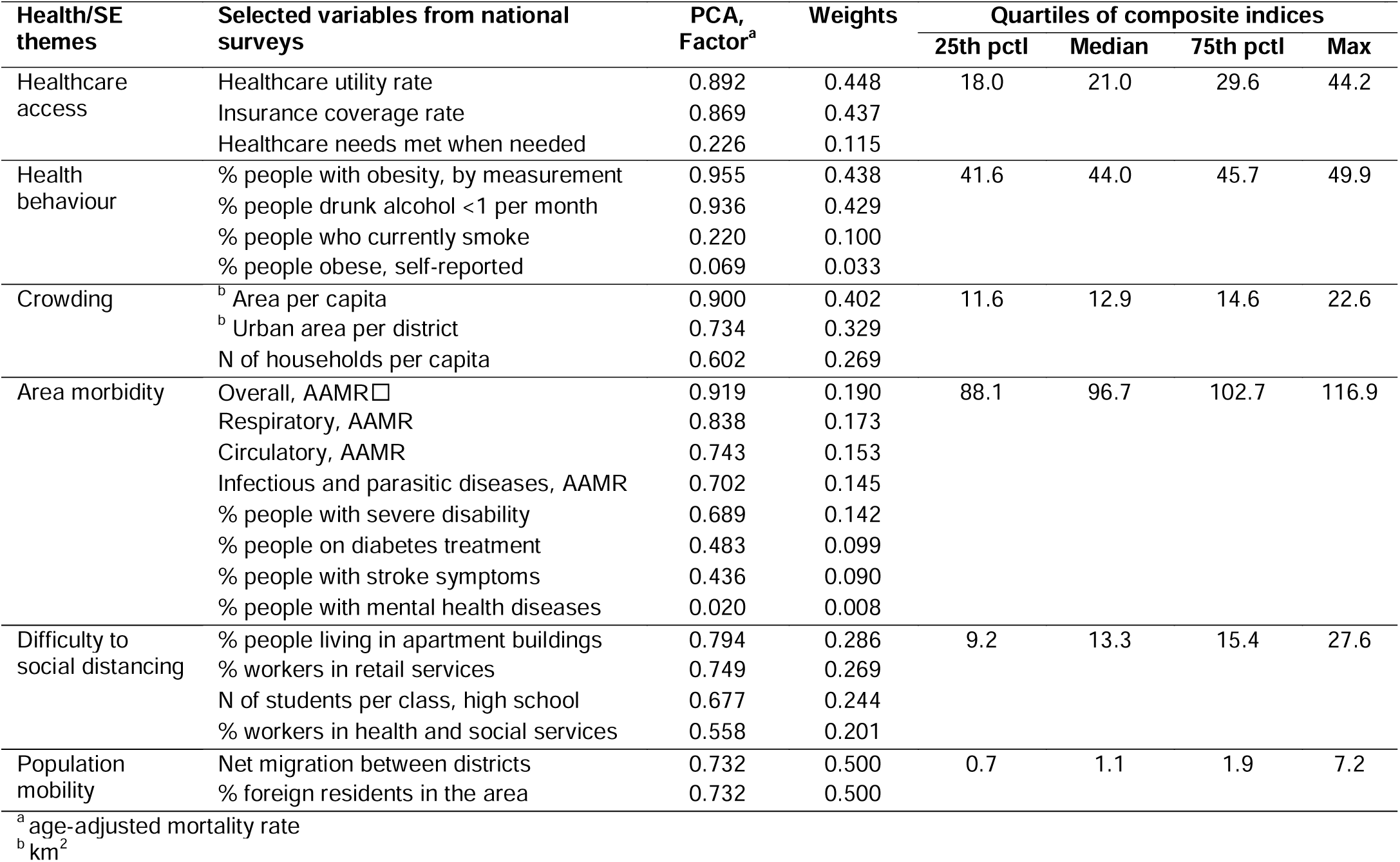
PCA details showing factor scores with their weights, and selected area health and SES variables and thematic composite indices

| Health/SE themes | Selected variables from national surveys | PCA, Factor<sup>a</sup> | Weights | Quartiles of composite indices |  |  |  |
| --- | --- | --- | --- | --- | --- | --- | --- |
|  |  |  |  | 25th pctl | Median | 75th pctl | Max |
| Healthcare access | Healthcare utility rate | 0.892 | 0.448 | 18.0 | 21.0 | 29.6 | 44.2 |
|  | Insurance coverage rate | 0.869 | 0.437 |  |  |  |  |
|  | Healthcare needs met when needed | 0.226 | 0.115 |  |  |  |  |
| Health behaviour | % people with obesity, by measurement | 0.955 | 0.438 | 41.6 | 44.0 | 45.7 | 49.9 |
|  | % people drunk alcohol <1 per month | 0.936 | 0.429 |  |  |  |  |
|  | % people who currently smoke | 0.220 | 0.100 |  |  |  |  |
|  | % people obese, self-reported | 0.069 | 0.033 |  |  |  |  |
| Crowding | <sup>b</sup> Area per capita | 0.900 | 0.402 | 11.6 | 12.9 | 14.6 | 22.6 |
|  | <sup>b</sup> Urban area per district | 0.734 | 0.329 |  |  |  |  |
|  | N of households per capita | 0.602 | 0.269 |  |  |  |  |
| Area morbidity | Overall, AAMR□ | 0.919 | 0.190 | 88.1 | 96.7 | 102.7 | 116.9 |
|  | Respiratory, AAMR | 0.838 | 0.173 |  |  |  |  |
|  | Circulatory, AAMR | 0.743 | 0.153 |  |  |  |  |
|  | Infectious and parasitic diseases, AAMR | 0.702 | 0.145 |  |  |  |  |
|  | % people with severe disability | 0.689 | 0.142 |  |  |  |  |
|  | % people on diabetes treatment | 0.483 | 0.099 |  |  |  |  |
|  | % people with stroke symptoms | 0.436 | 0.090 |  |  |  |  |
|  | % people with mental health diseases | 0.020 | 0.008 |  |  |  |  |
| Difficulty to social distancing | % people living in apartment buildings | 0.794 | 0.286 | 9.2 | 13.3 | 15.4 | 27.6 |
|  | % workers in retail services | 0.749 | 0.269 |  |  |  |  |
|  | N of students per class, high school | 0.677 | 0.244 |  |  |  |  |
|  | % workers in health and social services | 0.558 | 0.201 |  |  |  |  |
| Population mobility | Net migration between districts | 0.732 | 0.500 | 0.7 | 1.1 | 1.9 | 7.2 |
|  | % foreign residents in the area | 0.732 | 0.500 |  |  |  |  |

<sup>a</sup> age-adjusted mortality rate
<sup>b</sup> km<sup>2</sup>

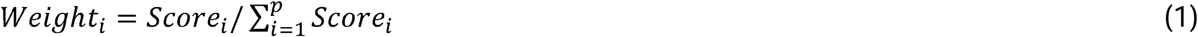

where *i* relates to each theme’s variable and *p* is the number of each theme’s variables. Each thematic composite index was computed as the weighted average for all 250 district values. For example, the composite index for *health behaviour* was calculated as:

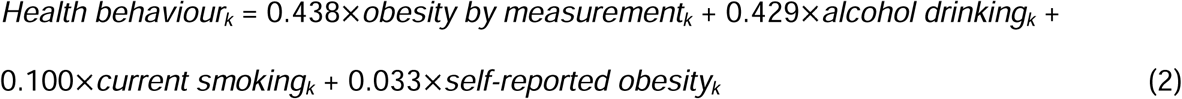

*Health behaviour*_*k*_ = 0.438 *obesity by measurement*_*k*_ + 0.429 *alcohol drinking*_*k*_ + where *k* is the original variable’s value for district *k*. Note that weights sum to 1 (0.438 + 0.429 + 0.100 + 0.033 = 1). Six thematic composite indices and an individual proxy for education (percentage of high school educated people) were used in the final models as independent variables.

The ratio of the 25^th^ percentile and each theme’s maximum value was for *healthcare access*: 2.5; *health behaviour*: 1.2; *crowding*: 1.9; *area morbidity*: 1.3; *education*: 1.5; *difficulty to social distancing*: 3.0; and *population mobility*: 10.3.

### Statistics

Our model outcome was the confirmed case counts of COVID-19 aggregated by 250 districts. Global negative binomial regression (GNBR) and GWNBR^7^ computed relative risk of COVID-19 associated seven area health/SE themes.

### Global models

GNBR models calculated relative COVID-19 risk for the entire study period and each pandemic phases. The global model was set as:

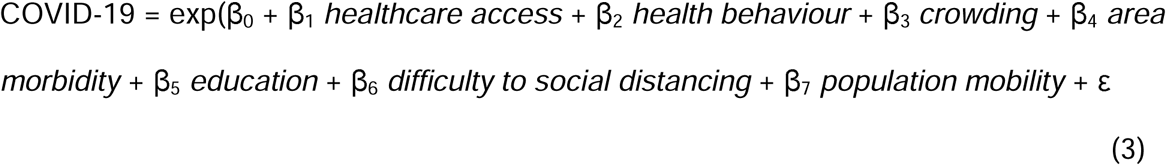

where, β_0_, …, β_n_ were the intercept and regression coefficients, whereas ε was the model random error.

### Local spatial models

We used Gaussian GWNBR to model a discrete count data and handle overdispersion issue. GWNBR computed parameter estimates for all districts following:

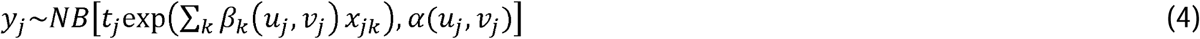

where (u_j_, v_j_) are the locations (coordinates) of the data points *j*, for *j* = 1, …, *n*. The models empirically computed bandwidth, via the cross-validation criterion, and achieved minimal Akaike information criterion (AIC) as:

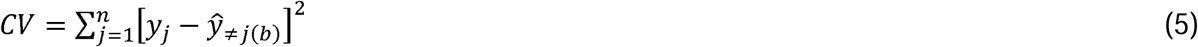

where ŷ_≠j(b)_ is the estimated value for point *j*, omitting the observation *j*, and *b* is the bandwidth. The likelihood of false-positives was corrected by the method of da Silva and Fotheringham^20^.

All statistical analyses including specific macro programs for spatial weight matrices and GWNBR models were implemented using SAS (version 9.4). Missing data (2%) were excluded from the analyses.

## Results

Figure 2 compared the spatial COVID-19 distribution across pandemic phases. The initial outbreak wave occurred in Daegu which then spread to Gyeongsangbuk-do and surrounding provinces in the early phase^16^. The second wave occurred in Seoul and its surrounding metropolises, Ulsan and Busan, and Gyeonggi-do province in the late phase of the pandemic.

**Figure 2.**
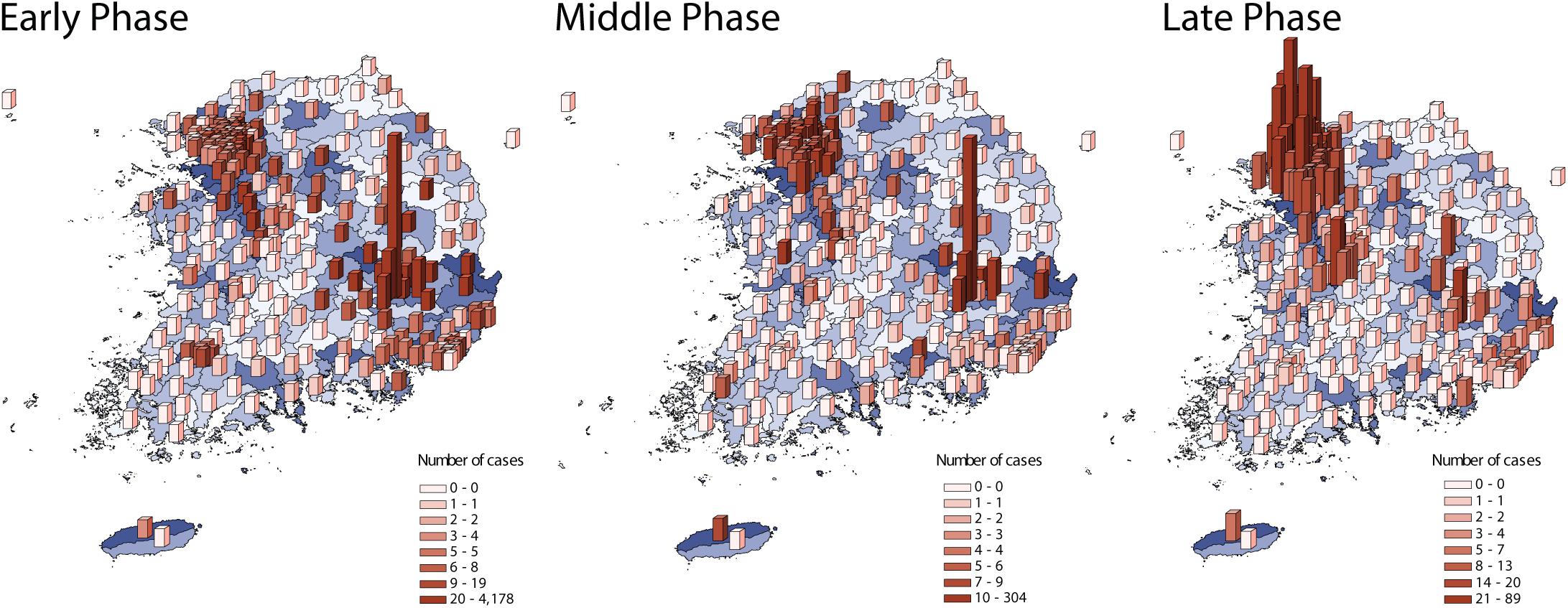
The spatial distribution of COVID-19 cases across pandemic phases. Early phase: from January 20 to March 20, 2020. Middle phase: March 21 to April 15, 2020. Late phase: April 16 to July 1, 2020. The background map in blue shades indicates the spatial variation of the population density. A blue colour gradient corresponds with a higher (darker) to lower (lighter) population density.

### SES measurement

Table 1 presents the PCA results including the contributing variables for each of the six composite indexes. The factor scores and weights of each contributing variable associated with the first PCA-identified component and the quartiles of the health/SE themes are shown.

### Global and local spatial models

In the entire study period model using GNBR, the COVID-19 risk associated with increased risky *health behaviour, area morbidity*, and *difficulty to social distancing* (Table 2). Inverse associations indicate an increased COVID-19 risk with reduced *healthcare access*, lower *education*, and increased outflux in *population mobility*. No substantial risk was associated with *crowding*.

**Table 2.**
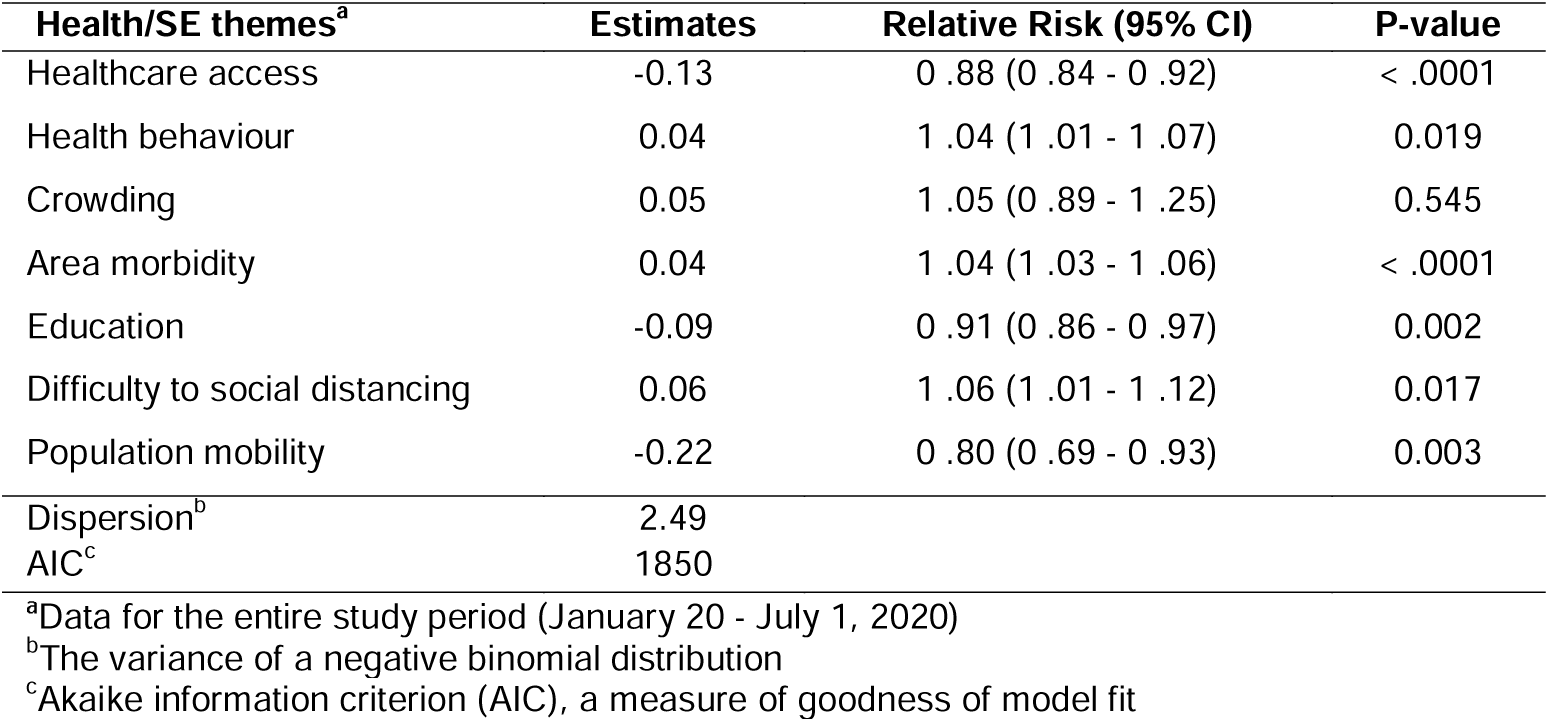
Parameter estimates and 95% CI of the relative risk of COVID-19 associated with health and SES determinants during the entire study period

We implemented global and local spatial models for the early, middle, and late pandemic phases. Figure 3 presents the relative COVID-19 risk with its 95% CI from GNBR models, and Figure 4, the relative risk spatial distribution from GWNBR associated with seven thematic areas by pandemic phases. Supplementary Table S4 provides more details on the stratified GNBR models. GWNBR fit data better than the global model given smaller AIC for the middle and late phases, respectively, (AIC_gwnbr_ ∼1034 vs AIC_gnbr_∼1044, AIC_gwnbr_ ∼1038 vs. AIC_gnbr_∼1074) except for the early phase of the pandemic (AIC_gwnbr_ ∼3533 vs AIC_gnbr_∼1527). This reflects the large spatial cluster emerging from Daegu church^16, 21^ activities during the early phase that subsequently spread to its neighbouring districts. The GNBR and GWNBR model results agreed across all pandemic phases. In the early phase, lower *healthcare access* and *education*, and increased risky *health behaviour, area morbidity, difficulty to social distancing*, and *population mobility* associated with higher COVID-19 risk. The *crowding-associated* risk was not significant in GNBR. In the middle phase, *healthcare access, area morbidity, education, and difficulty to social distancing* remained important risk determinants. In the late phase, only *healthcare access, health behaviour*, and *increased crowding* significantly determined the COVID-19 risk.

**Figure 3.**
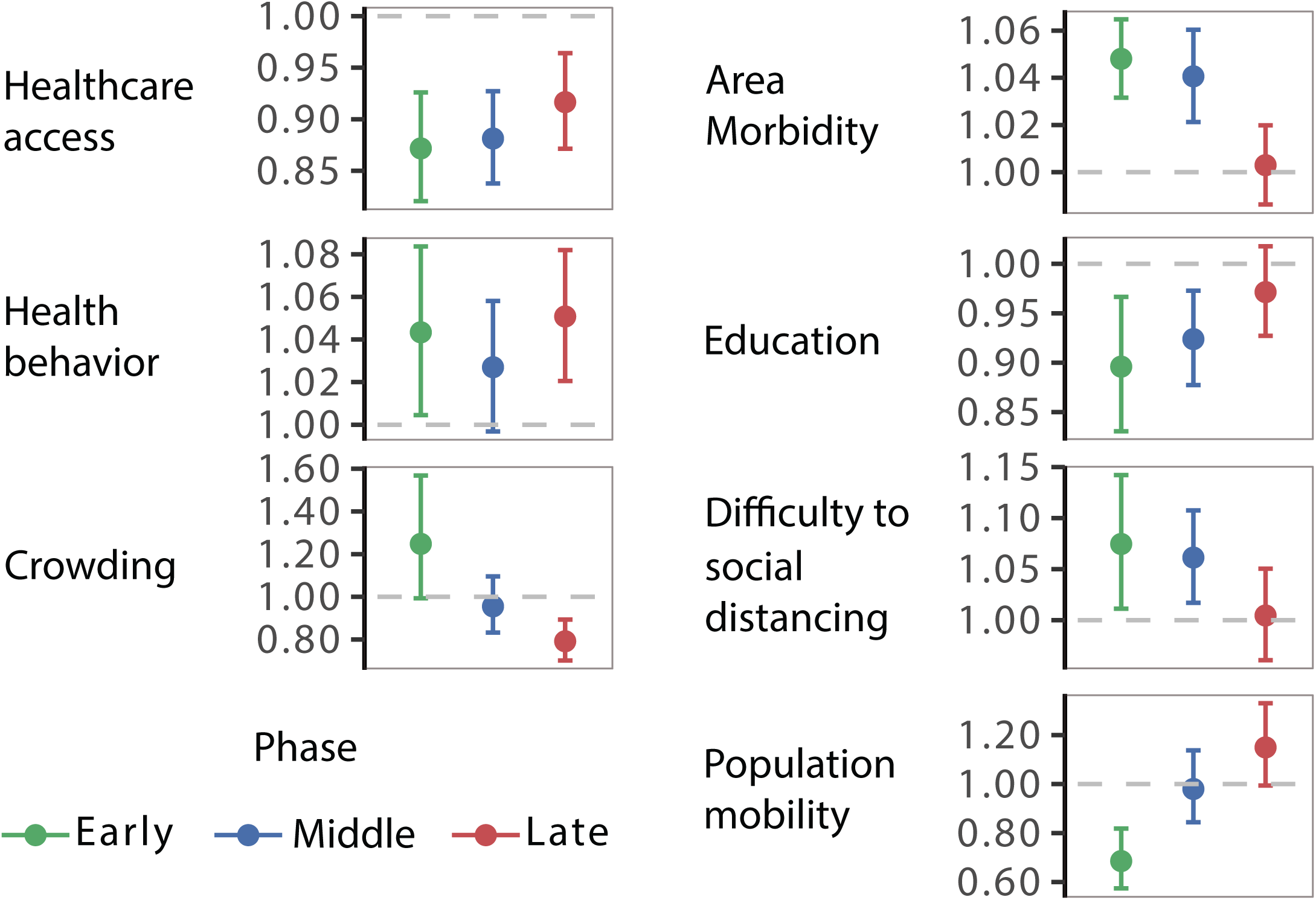
Relative Risk of COVID-19 associated with area health and SES determinants. Each panel shows the relative risk and its 95% confidence intervals associated with each thematic area. Colours represent the pandemic’s early (January 20 to March 20, 2020), middle (March 21 to April 15, 2020), and late phases (April 16 to July 1, 2020). The dashed line shows the reference level (1). Values over or below the reference line indicate statistically significant results at α = 0.05.

**Figure 4.**
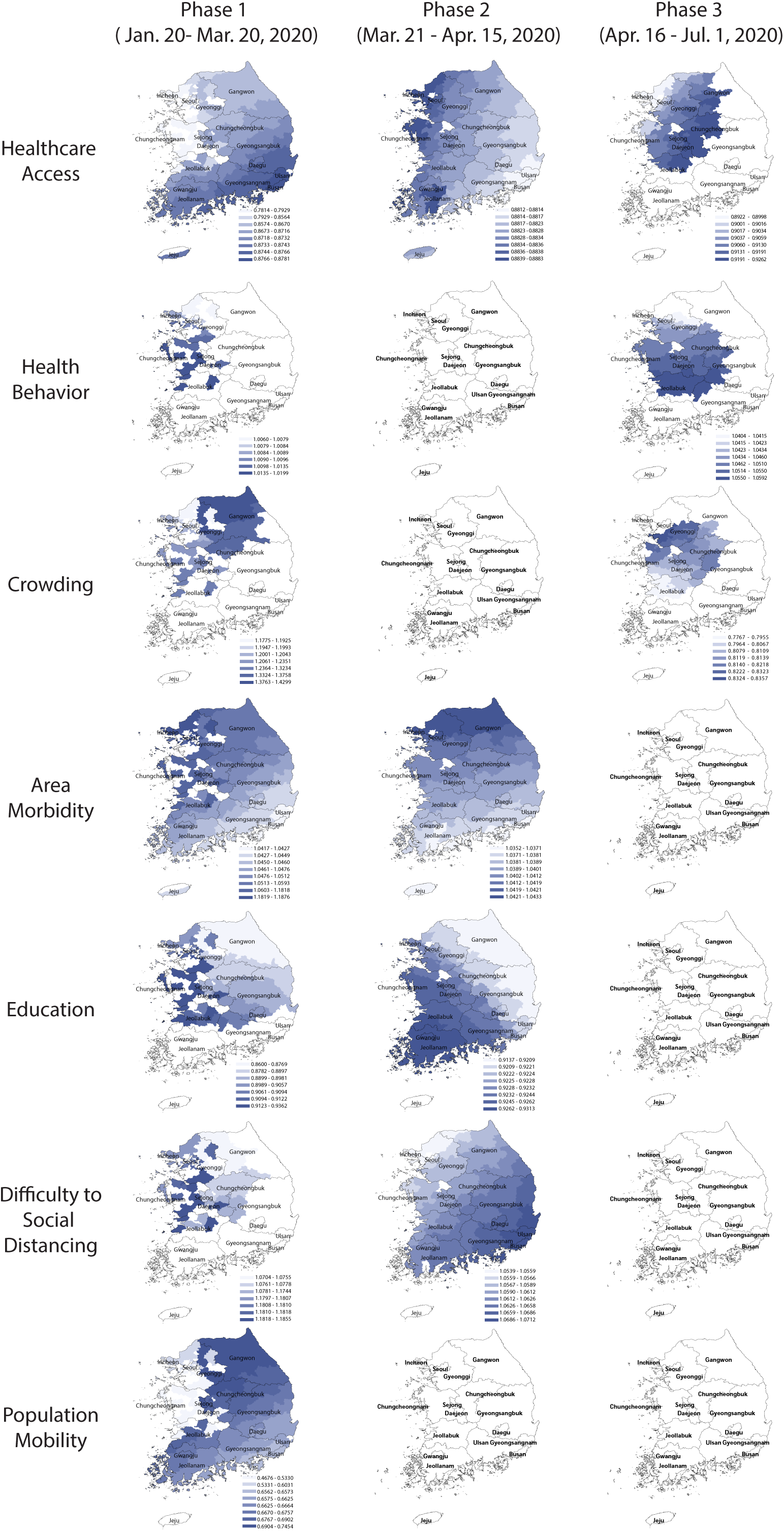
Spatial variation in the relative risk of COVID-19 associated with area-health and SES themes in the early, middle, and late phases of the pandemic, GWNBR models. In the maps, the blue colour gradient corresponds with larger (darker) to lower (lighter) relative risk. Areas in white indicate the relative risks are statistically not significant (α = 0.05). Columns titled Early, Middle, and Late phases refer to the pandemic’s early (January 20 to March 20, 2020), middle (March 21 to April 15, 2020), and late phases (April 16 to July 1, 2020).

GWNBR created early phase maps showing higher risk in non-contiguous districts. This higher risk reflected virus transmission in the initially affected districts before spreading over expanded areas.

During early phase, we found protective effect of improved healthcare access, higher education and outbound population mobility, whereas, the disease risk was increased in the districts with higher risky health behaviour, area morbidity rate and difficulty to social distancing.

In the middle phase, only healthcare access, area morbidity rate, education and difficulty to social distancing remained as the key risk-determinants with the same directions but reduced strengths. Spatial shifts from the early phase was from the northwest toward the capital and southwest regions.

In the late phase, healthcare access, risky healthy behaviour and area crowding were primary risk-determinants. We observed protective effect of improved healthcare access while risky health behaviour was the significant risk factor. In contrast to the early phase, we found higher disease risk in more crowded districts. Overall spatial pattern of the later phase of the pandemic was concentrated around the capital and the middle regions. Taken the type of risk-determinants and their spatial distributions across the three phases together, our results showed that pandemic had evolved from lower to higher density areas which led to the second wave that emerged in Seoul and its surrounding areas.

We observed noticeable spatial shifts in the risk determinants over the study period. *Difficulty to social distancing* increased COVID-19 risk in the capital and middle regions in the early phase which then shifted to the country’s southeast part in the middle phase. *Area morbidity*-associated risk was concentrated in the western part which then gradually shifted north in the middle phase. *Education-*associated risk was higher in the west in the early phase until it shifted southwest in the middle phase. *Population mobility* elevated COVID-19 risk only in the early phase for South Korea’s northern, eastern, and western parts.

We investigated the correlations between all pairs of composite indices (Table S5). The largest Pearson’s r was 0.603 between *healthcare access* and *area morbidity*. We verified no multicollinearity given that the model’s standard error of both *healthcare access* (0.025) and *crowding* (0.09) were small. For a direct comparison between non-spatial and spatial models, GNBR and GWNBR were carried out with the same variables and stratified by the same periods (NBR: Figure 3, and GWNBR: Figure 4). AIC and dispersion coefficients were used to compare the models’ goodness of fit.

## Discussion

This study provided a methodology to map COVID-19 risk associated with multiple SES and pandemic-specific factors with high geographical granularity. The simultaneous influence of multiple SES determinants defines disease risk. By creating multiple composite SES indexes using PCA, this study clarified whether certain SES determinants independently contributed to the COVID-19 risk over and above other SES factors. The social theories-enhanced SES measurement in our refined model, CBF, allowed us to conceptualize the mechanistic pathway from the macro-level domains (material, human, and capital) through the SES variable selection process. Guided by the conceptual framework, we identified COVID19-specific determinants that cause differential exposure, susceptibility, and disease severity.

GWNBR created a continuous surface of relative COVID-19 risk for all 250 districts associated with area-health and socioeconomic determinants by the pandemic phases (Figure 4). Our findings are consistent with individual and population-level studies that reported elevated COVID-19 risk associated with less *healthcare access*^22^, and *education*^23, 24^, and more *risky health behaviour, crowding, specific comorbidities*^13, 14, 17^, *difficulty to social distancing*^15, 25^ and *population mobility*^26^. Our study’s high internal validity was shown since the GNBR and GWNBR results agreed except for *crowding* in the early phase.

Our approach captured statistically and noticeably high spatial variation by pandemic phases for all themes, consistent with the reported pattern of COVID-19 distribution in the country^16, 21^. Since its first confirmed case on January 20th, 2020, South Korea experienced two major outbreak waves in Daegu and Seoul, and the surrounding Gyeonggi-do province, respectively, in February (early phase) and May 2020 (late phase). The country responded to the first wave with nationwide directives that included mass testing based on contact tracing, self-quarantine/isolation, strengthening medical centres for rapid diagnostics, emergency medical responses, and treatment aids^16^. These specific measures combined with high public adherence to the school and business closures, personal hygiene, and social distancing significantly dropped the case counts until the second wave erupted in May when businesses reopened^21^.

SES-related risk declined over the pandemic phases. Analysis stratified by periodic phases found that the initially high risk in the early period gradually decreased in strength and spatial coverage in the middle and late phases (Figure 3 & Figure 4). The exceptions were health behaviour and crowding-associated risk, which increased in the late phase. Reductions could be explained by the impact of effective control measures that lowered the risk associated with these determinants and/or a drop in an effective reproduction number (Re), as the number of infection-susceptible people decreased over time^27^.

*Types of risk determinants for SES-related COVID-19 risk varied over the pandemic phases*. In the early phase, all health/SE themes were key risk factors. In the middle phase, all of the previous risk factors except for risky *health behaviours, population mobility*, and *crowding* were high. In the late phase, only increased risky *health behaviour*, increased *crowding* and reduced *healthcare access* remained directly associated with COVID-19 incidence. The middle phase’s lessened risk associated with risky *health behaviours, population mobility* and *crowding* could reflect the impact of the Prime Minister’s declaration. This implemented active interventions for social distancing, community health education, testing with local contact tracing and screening centre follow-ups, and quarantine and isolation during March’s first weeks. Notably, the late phase findings are consistent with the risk factors reported associated with the second wave in early May. During our study’s late phase, South Korea scaled up testing and treatment in its existing health care centres^21, 28^ which may have made improved *healthcare access* a key measure for combating COVID-19. Our finding of *healthcare access* exerting a stronger protective effect in the late phase rather than the earlier phases supports this. In addition, our findings may have captured behavioural fatigue at a population-scale in response to the country’s multiple quarantine period extensions^29^. Behavioural fatigue was observed in other countries that likely were exacerbated by business re-openings in early May. Elevated risk associated with increased *crowding* during the outbreak’s second wave, occurred in South Korea’s most crowded region: Seoul and its surroundings. However, in the early pandemic, elevated risk associated with decreased crowding (larger area per person) in the most sparsely populated districts (excluding Jeju-island) such as Gangwon-do, Chungcheongbuk-do and specific cities in Gyeonggi-do. It’s likely that one specific mass gathering at a Daegu church^16, 21^ was a more relevant risk factor than general crowding, supported by Daegu’s lack of crowding-associated risk. Figure 4 also shows that population-mobility and difficulty to social distancing were important risk determinants in the pandemic’s early phase until these areas were regulated by public health interventions during the later phases.

The increase in health behaviour-associated risk is consistent with reports showing greater risk with poor *emotional health*^23^, *smoking*^30^, and *obesity*^14, 31^. Lusignan et al.,2020^14^ reported smoking was a protective factor; however, the authors warned that the low proportion of current smokers in their study sample (11.4%), resulted in a wide confidence interval of the reported odds ratio, 0.59 (0.42-0.83). This increases the uncertainty of their result. Greater COVID-19 risk among the people with lower education has been explained as a reduced *awareness of disease risk* and *low-income*. Thus, the inability to afford unemployment resulted in the reduced exertion of protective measures^23, 24^.

*Spatial variation in the SES-related risk factors across the pandemic phases potentially reflect the geography-specific control measures and/or the differential public response to the measures*. GWNBR models revealed the pandemic phase-specific spatial variation for all health/SE themes except for population mobility which was not significant beyond the early phase. This may indicate that the effectiveness of the control measures varied over time potentially due to differential interventions or public response across the municipal districts. Our findings may also indicate a dynamic change in population vulnerability throughout the pandemic “a person not considered vulnerable at the outset of a pandemic can become vulnerable depending on the policy response” as a *Lancet* editorial stated^32^.

The factors increasing our recommended framework’s robustness include: 1) SES measurement and relationship conceptualization of the exposure (health/SE themes) and outcome (COVID-19 incidence) based on the refined conceptual framework; 2) joint use of conceptual and statistical modelling; 3) complementary use of global and local spatial statistics; and 4) stratified analysis by pandemic phases that enable us to capture the spatial variation over pandemic phases. However, this methodological framework relies on carefully collected country-specific data.

Biases due to differential testing rates country-wide are low since the testing was based on contact tracing, and government-supported (free). Our study is subject to ecological fallacy inherent to the study design. However, our empty hierarchical mixed model accounting for the individual and district-level data shows that 61% of the COVID-19 incidence distribution variation was explained by the district-level factors, leaving 39% of the variability for an explanation by individual factors. We verified that the data estimation for Daegu city subparts did not affect the study results. The comparison of the intercept, standard error, relative risk and P-value between the models with and without the estimated data showed that the intercept and standard error were diminished by 2.2% and by 9.2%, respectively in the models including estimated data [each calculated by 100 (−12.29 -(−12.56))/-12.29 and 100 (1.88-2.95)/1.88, respectively]. A significance level change was observed for none of the model estimates, except for *crowding*. The P-value changed from ∼0.06 to ∼0.04 when the estimated data were excluded. However, the *crowding-associated* risk remains significant at P = 0.1. Model details are provided in Table S2. To assess the periodic trend in the relative COVID-19 risk associated with SES factors, we conducted stratified analyses by the early, middle, and late phases corresponding with January 20-March 20, March 21-April 15, and April 16-July 1, 2020.

We intended this framework to improve international knowledge exchange and enable rapid pandemic responses in high-risk populations. To illustrate practical application of this framework in South Korea, decision-makers could have prioritized improved healthcare access and promoted protective health behaviours focusing more on the crowded areas in the capital and surrounding regions in the second wave of the pandemic. Such precision targeting can bolster preventive measures to reduce the healthcare burden and economic damage.

## Data Availability

The data and SAS code underlying this article will be shared on reasonable request to the corresponding author.

## Acknowledgments

The work described here was not funded by any source. Dr. Kouzoukas receives research grant support from the US Department of Veterans Affairs (I21 RX003170 to DEK). The views expressed here are those of the authors and do not necessarily reflect that of any government agency or institution.

## Supplementary tables

**Table S1.** Confirmed cases of COVID-19 diagnosed during January 20 - July 1, 2020 in South Korea by pandemic phases

| Province | Phase 1 | Phase 2 | Phase 3 |
| --- | --- | --- | --- |
| Busan | 99 | 18 | 24 |
| Chungcheongbuk | 34 | 11 | 12 |
| Chungcheongnam | 85 | 17 | 28 |
| Daegu | 6358 | 529 | 35 |
| Daeguon | 21 | 17 | 80 |
| Gangwon | 30 | 20 | 13 |
| Gwangju | 48 | 2 | 3 |
| Gyeonggi | 324 | 325 | 567 |
| Gyeongsangbuk | 1098 | 111 | 45 |
| Gyeongsangnam | 86 | 26 | 17 |
| Incheon | 38 | 47 | 253 |
| Jeju | 3 | 9 | 6 |
| Jeollabuk | 8 | 9 | 4 |
| Jeollanam | 4 | 10 | 0 |
| Sejong | 34 | 10 | 7 |
| Seoul | 298 | 278 | 656 |
| Ulsan | 32 | 8 | 15 |

**Table S2.** GNBR model estimates with and without the estimated data for Daegu’s subparts

| Included estimated data |  |  |  |  | Excluded estimated data |  |  |  |
| --- | --- | --- | --- | --- | --- | --- | --- | --- |
| Health/SE themes<sup>a</sup> | Estimate<sup>b</sup> | SE | RR (LCL - UCL) | P-value | Estimate | SE | RR (LCL - UCL) | P-value |
| Area-morbidity | 0.05 | 0.01 | 1.05 (1.03 - 1.06) | <.0001 | 0.04 | 0.01 | 1.04 (1.03 - 1.06) | <.0001 |
| Education | -0.11 | 0.04 | 0.90 (0.83 - 0.97) | 0.005 | -0.12 | 0.04 | 0.89 (0.82 - 0.96) | 0.003 |
| Crowding | 0.22 | 0.12 | 1.25 (0.99 - 1.57) | 0.06 | 0.27 | 0.13 | 1.30 (1.01 - 1.69) | 0.04 |
| Difficulty to social distancing | 0.07 | 0.03 | 1.07 (1.01 - 1.14) | 0.02 | 0.08 | 0.03 | 1.08 (1.02 - 1.15) | 0.01 |
| Population mobility | -0.38 | 0.09 | 0.69 (0.57 - 0.82) | <.0001 | -0.34 | 0.09 | 0.71 (0.59 - 0.85) | 0.0002 |
| Healthcare access | -0.14 | 0.03 | 0.87 (0.82 - 0.93) | <.0001 | -0.13 | 0.03 | 0.88 (0.82 - 0.93) | <.0001 |
| Health behaviour | 0.04 | 0.02 | 1.04 (1.00 - 1.08) | 0.03 | 0.04 | 0.02 | 1.04 (1.00 - 1.08) | 0.05 |
| Dispersion | 3.68 |  |  |  | 3.69 |  |  |  |
| AIC | 1527 |  |  |  | 1425 |  |  |  |

<sup>a</sup>Study period: from January 20 through July 1, 2020
<sup>b</sup>Parameter estimates; SE: Standard error; RR: Relative Risk; LCL: lower boundary of 95% confidence interval; UCL: upper boundary of 95% confidence interval.

**Table S3.** Data sources reviewed and used for SES measurement

| Data sources | Data items |
| --- | --- |
| Korean Community Health Survey 2018, Korea Centres for Disease Control and Prevention | (%) people with obesity, by measurement<br>(%) people drunk alcohol <1 per month<br>(%) people who currently smoke<br>(%) people obese, self-reported<br>(%) of people who used health care last year<br>(%) people who could not use healthcare when needed last year<br>(%) people with depression based of PHQ-9<sup>a</sup> screening<br>(%) of the person who is all aligned for the early symptoms of stroke<br>(%) of the person on insulin and other treatment specific to diabetes mellitus |
| Health Insurance Statistics 2018, National Health Insurance Corporation | (%) people with health insurance |
| Disability Status 2018, Ministry of Health and Welfare | (%) people with severe disability |
| Death Cause Statistics 2018, National Statistics Agency | Age adjusted mortality rate due to neoplasm<br>Age adjusted mortality rate due to circulatory system disease<br>Age-adjusted mortality rate from infectious parasitic diseases<br>Overall, age-adjusted mortality rate<br>Age adjusted mortality rate due to respiratory diseases |
| Korean Census Bureau 2015 | (%) people with high school education<br>(%) of foreign registered people<br>Number of people per household |
| Office of Statistics 2015, Regional Statistics | GDP per capita in million won |
| Internal Migration Statistics 2018, Statistics Korea | Internal net migration between regions |
| State of Urban Planning 2018, Ministry of Land, Infrastructure, Transport and Tourism | Area per capita<br>Urban area per capita |

<sup>a</sup>PHQ-9: Patient Health Questionnaire -9, standard survey tool

**Table S4.** Parameter estimates and the Relative Risk of the COVID-19 incidence associated with health and SES determinants by three time periods corresponding with the early, middle and late phases

|  | Early phase |  |  | Middle phase |  |  | Late phase |  |  |
| --- | --- | --- | --- | --- | --- | --- | --- | --- | --- |
|  | Estimates | RR (LCL - UCL) | P-value | Estimates | RR (LCL - UCL) | P-value | Estimates | RR (LCL - UCL) | P-value |
| Healthcare access | -0.14 | 0.87 (0.82 - 0.93) | <.0001 | -0.13 | 0.88 (0.84 - 0.93) | <.0001 | -0.09 | 0.92 (0.87 - 0.96) | 0.001 |
| Health behaviour | 0.04 | 1.04 (1.00 - 1.08) | 0.028 | 0.03 | 1.03 (1.00 - 1.06) | 0.080 | 0.05 | 1.05 (1.02 - 1.08) | 0.001 |
| Crowding | 0.22 | 1.25 (0.99 - 1.57) | 0.058 | -0.05 | 0.96 (0.83 - 1.10) | 0.512 | -0.23 | 0.79 (0.70 - 0.89) | 0.000 |
| Area morbidity | 0.05 | 1.05 (1.03 - 1.06) | <.0001 | 0.04 | 1.04 (1.02 - 1.06) | <.0001 | 0.003 | 1.00 (0.99 - 1.02) | 0.729 |
| Education | -0.11 | 0.90 (0.83 - 0.97) | 0.005 | -0.08 | 0.92 (0.88 - 0.97) | 0.003 | -0.03 | 0.97 (0.93 - 1.02) | 0.223 |
| Difficulty to social distancing | 0.07 | 1.07 (1.01 - 1.14) | 0.021 | 0.06 | 1.06 (1.02 - 1.11) | 0.006 | 0.005 | 1.00 (0.96 - 1.05) | 0.843 |
| Population mobility | -0.38 | 0.69 (0.57 - 0.82) | <.0001 | -0.02 | 0.98 (0.84 - 1.14) | 0.791 | 0.14 | 1.15 (0.99 - 1.33) | 0.061 |

Early phase: January 20 to March 20, 2020; Middle phase: March 21 to April 15, 2020; Late phase: April 16 to July 1, 2020 RR: Relative Risk; LCL: lower boundary of 95% confidence interval; UCL: upper boundary of 95% confidence interval.

**Table S5.** Matrix table of Pearson’s correlation coefficients (r)

|  | **Healthcare access** | **Health behaviour** | **Crowding** | **Area morbidity** | **Education** | **Difficulty to social distancing** | **Population mobility** |
| --- | --- | --- | --- | --- | --- | --- | --- |
| Healthcare access | 1 |  |  |  |  |  |  |
| Health behaviour | 0.547 | 1 |  |  |  |  |  |
| Crowding | 0.568 | 0.124 | 1 |  |  |  |  |
| Area morbidity | 0.604 | 0.534 | 0.415 | 1 |  |  |  |
| Education | 0.127 | 0.390 | -0.080 | 0.510 | 1 |  |  |
| Difficulty to social distancing | -0.032 | 0.370 | -0.047 | 0.311 | 0.315 | 1 |  |
| Population mobility | -0.096 | 0.086 | -0.019 | 0.018 | 0.163 | 0.129 | 1 |

